# Site-Specific Cancer Incidence among Clinical Subtypes of Newly Diagnosed Type 2 Diabetes in the United States

**DOI:** 10.64898/2026.08.17.26360595

**Authors:** Zhongyu Li, Chang Liu, Mary Beth Weber, Mohammed K. Ali, Craig Hofmeister, Jithin Sam Varghese

## Abstract

**Background:** Type 2 diabetes (T2D) is associated with elevated rates of several cancers and is increasingly recognized as a heterogeneous disease, but whether its clinically distinct subtypes carry different cancer risks is unknown.

**Methods:** In this matched retrospective cohort study using electronic health record data from the Epic Cosmos Research Platform (2012-2025), adults with newly diagnosed T2D were classified into severe insulin-deficient (SIDD, 21.6%), mild obesity-related (MOD, 23.5%), mild age-related (MARD, 40.7%), or mixed (14.1%) subtypes using validated algorithms and matched to adults without diabetes on age, sex, and body mass index. Cause-specific Cox models estimated adjusted hazard ratios (HRs) for seven site-specific cancers, accounting for competing risks. Cancer screening uptake was assessed as a secondary outcome.

**Results:** Among 575,139 adults with T2D and 689,719 without diabetes (median follow-up, 3.8 years), MARD had the highest cancer incidence (17.3 per 1,000 person-years). Relative to adults without diabetes, rates of colorectal, pancreatic, liver, endometrial, and ovarian cancer were elevated across subtypes, with the highest hazards in SIDD (HR=3.87, 95% CI=3.51 to 4.27) and mixed phenotypes. Prostate cancer rates were lower in all subtypes, most markedly in MOD (HR=0.60, 95% CI=0.55 to 0.64). Rates of breast cancer were higher among mixed (HR=1.12, 95% CI=1.05 to 1.19) and lower among MOD (HR=0.85, 95% CI=0.80 to 0.90). Mammography and prostate-specific antigen screening were lower across subtypes.

**Conclusions:** Site-specific cancer incidence and screening uptake differed across clinically defined subtypes of T2D. Subtype classification from routine clinical data may inform targeted cancer surveillance, though further study is needed before clinical use.

## INTRODUCTION

Type 2 diabetes (T2D) affects more than 38 million adults in the United States and accounts for over 1.2 million new diagnoses each year.^1^ While cardiovascular mortality in T2D has declined over recent decades, cancer has become an increasingly important contributor to diabetes-related morbidity and mortality.^2,3^ Recent multinational evidence from 11 high-income jurisdictions showed that cancer is the leading cause of death among people with diabetes in four settings, accounting for more than 22% of diabetes related deaths.^4^ Characterizing which individuals with T2D carry the greatest risks for site-specific cancer is therefore increasingly relevant to cancer prevention and clinical care.

Current cancer prevention and screening in T2D largely follow the same age-, sex-, and obesity-based strategies used for the general population.^5,6^ This approach assumes relative homogeneity in cancer risk within T2D and may obscure clinically meaningful variation across individuals. Recent advances have shifted the understanding of T2D from a single disease to distinct metabolic subtypes characterized by differences in insulin resistance, β-cell dysfunction, obesity, lipid dysregulation, and aging.^7,8^ The most widely reproduced subtypes (Severe Insulin-Deficient Diabetes [SIDD], Severe Insulin-Resistant Diabetes [SIRD], Mild Obesity-related Diabetes [MOD], and Mild Age-related Diabetes [MARD]) show variations in clinical presentations, disease progression rates, and risks for vascular complications.^8–11^ For instance, SIDD is characterized by insulin deficiency and severe hyperglycemia at diagnosis, whereas SIRD is defined by pronounced insulin resistance and obesity and is associated with elevated risks of cardiovascular disease and metabolic dysfunction–associated steatotic liver disease.^7^ Importantly, these subtype-defining traits are also relevant to carcinogenesis. Hyperglycemia, hyperinsulinemia, adiposity, and lipid dysregulation have each been implicated in tumor initiation and progression,^12,13^ suggesting that cancer risk in T2D may also vary across metabolic phenotypes.

Evidence directly testing this hypothesis remains limited. Most prior studies have examined diabetes timing, duration, or individual metabolic traits rather than multidimensional clinical phenotypes,^14–18^ and few have evaluated site-specific cancer outcomes across T2D subtypes. To address these gaps, we conducted a retrospective cohort study using EHR data from the United States to evaluate site-specific cancer risk across clinically defined subtypes of newly diagnosed T2D, compared with adults without diabetes. By characterizing cancer risk across clinically observable T2D subtypes, this work aims to inform future risk stratification approaches and generate evidence relevant to precision cancer prevention in diabetes care.

## METHODS

### Data Source

We conducted a matched retrospective cohort study using data from the Epic Cosmos Research Platform, a HIPAA-compliant, de-identified electronic health record (EHR) database comprising longitudinal data on over 300 million unique patients from all 50 U.S. states and the District of Columbia.^19^ Patients are linked longitudinally across participating health systems through a privacy-preserving process and are broadly representative of the U.S. population.^20^

### Study Population

#### Adults with type 2 diabetes

We identified adults aged 18–99 years with newly diagnosed T2D between January 1, 2012, and December 31, 2023, using the Surveillance, Prevention, and Management of Diabetes Mellitus (SUPREME-DM) computable phenotype (see **Supplementary Methods**).^21^ The date of T2D diagnosis was defined as the date on which the second SUPREME-DM criterion was met. To ensure the availability of baseline covariates, we required evidence of healthcare utilization, defined as at least one inpatient or outpatient encounter in each of the two years preceding T2D diagnosis date for cohort entry. We excluded individuals with a history of any cancer prior to diagnosis.

#### Adults without diabetes

We matched individuals with T2D to those without diagnosed diabetes. We excluded individuals with any recorded diagnosis codes for diabetes (type 1 diabetes, type 2 diabetes, gestational diabetes, and other specified diabetes) from problem lists and billing records. We then randomly sampled 5% of the remaining population (approximately 13.5 million individuals) to improve computational efficiency. For each individual, we defined the eligibility date as the earliest date at which both age ≥35 years and BMI ≥25 kg/m² were recorded, aligning cohort entry with U.S. Preventive Services Task Force eligibility thresholds for diabetes screening among adults with overweight/obesity.^22^ We additionally restricted the analysis to those individuals with at least one in-person encounter in each of the two years following the eligibility date and excluded those with a history of cancer (see **Figure S1**). The index dates for T2D and those without diabetes were defined as the date of diagnosis and the eligibility date shifted forward by two years, respectively.

Adults without diabetes were matched to adults with newly diagnosed T2D using coarsened exact matching (CEM) on age at index date (5-year categories), sex, and BMI (18.5, 25.0, 30.0, 35.0, and 40.0 kg/m^2^) measured within one year of the index date (**Supplementary Methods**).^23^ The analytic sample included 1,264,858 individuals (689,719 without diabetes and 575,139 with type 2 diabetes) (**Figure 1**).

**Figure 1.**
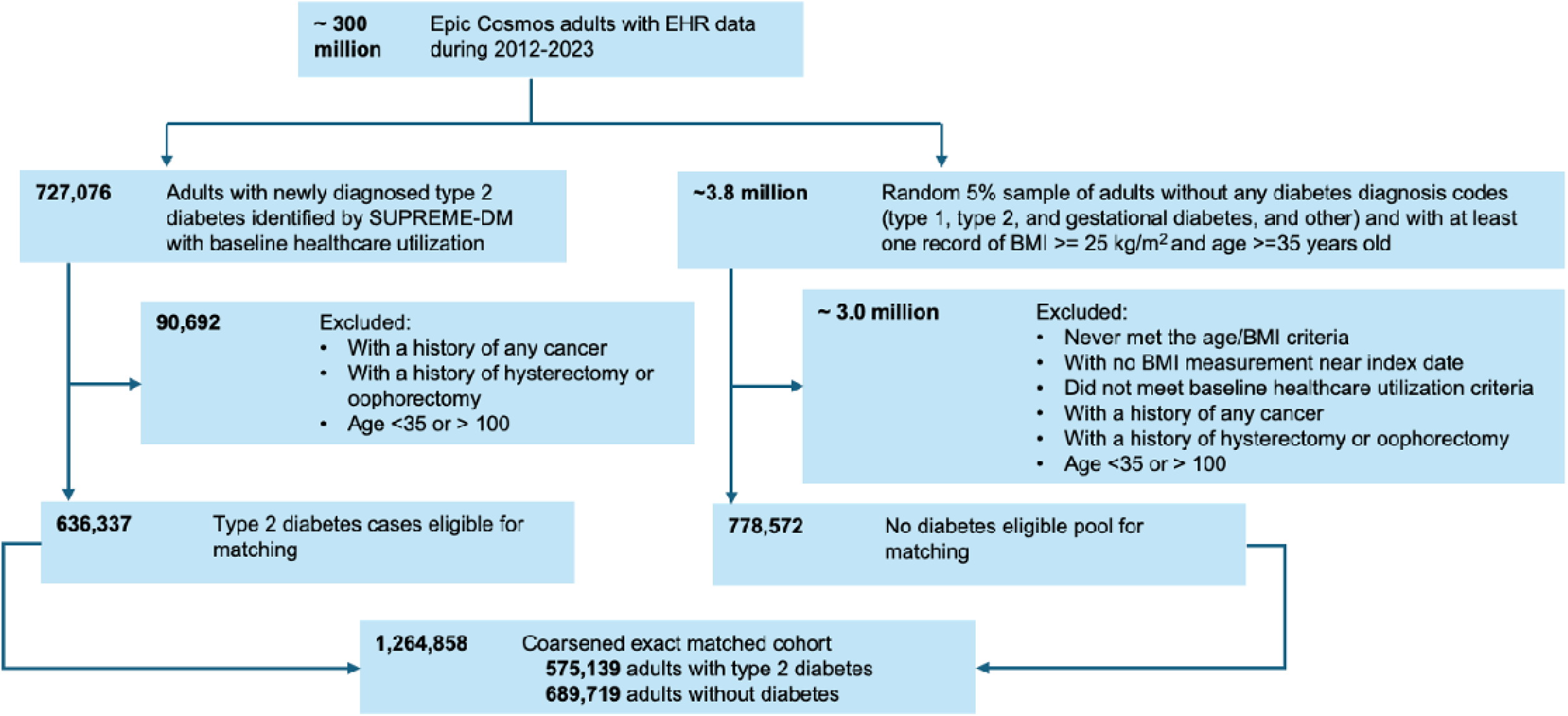
Participant selection flow diagram

### Variables

Adults with newly diagnosed T2D were classified into four subtypes using a previously described EHR-based classification approach that operationalizes adult-onset T2D subtypes using routinely available clinical variables (e.g., age at diagnosis, body mass index, HbA1c, blood pressure, and lipid measures).^11^ Breily, individuals were assigned sequentially to severe insulin-deficient diabetes (SIDD), mild age-related diabetes (MARD), and mild obesity-related diabetes (MOD); those not meeting criteria for these subtypes were categorized as Mixed. Classification model discrimination and assignment details are provided in the **Supplementary Methods**.

#### Primary outcome

Incidence of site-specific cancers (colorectal, prostate, pancreatic, liver, breast, endometrial, and ovarian) within 10 years after the index date was identified using International Classification of Diseases, Tenth Revision, Clinical Modification (ICD-10-CM) diagnosis codes recorded in Epic Cosmos (**Table S1**). Follow-up accrued from the index date until incident cancer diagnosis or the last recorded in-person encounter before the end of the study period (December 31, 2025), whichever occurred first.

#### Secondary outcome

As a secondary analysis undertaken to help interpret the observed cancer incidence patterns, we assessed cancer screening uptake across subtypes. Screening events, including mammography, prostate-specific antigen (PSA) testing, and colorectal cancer screening, were identified using Current Procedural Terminology (CPT) codes (**Table S1**), and screening uptake was defined as the time from the index date to the first post-index screening event.

### Statistical analysis

Covariate balance in the matched cohort was assessed using weighted standardized mean differences (SMDs), with absolute values less than 0.1 considered indicative of adequate balance. Cause-specific Cox proportional hazards models were used to estimate hazard ratios (HRs) and 95% confidence intervals (CIs) for the associations of T2D subtypes and no diabetes with site-specific cancers.^24^ All models adjusted for prespecified covariates: age, sex, race and ethnicity, smoking status and alcohol use within one year of the index date, body-mass index at or nearest the index date, year of cohort entry, age-adjusted Charlson Comorbidity Index (CCI) ^25^ in the prior five years, pre-index healthcare utilization in the prior two years (total encounters, number of months with ≥1 encounter, and mean encounters per active month), cardiometabolic medication use in the prior year, and cancer site-specific covariates selected a priori in the prior five years (**Table S2**). Similarly, in the secondary analysis, we estimated differences in cancer screening uptake across subtypes using cause-specific Cox models for time to the first post-index mammography, PSA test, and colorectal cancer screening event, adjusted for the same covariate set.

We conducted several sensitivity analyses to evaluate the robustness of our findings. To reduce potential reverse causation and detection bias, we performed lag-time analyses excluding cancers diagnosed within the first two years after the index date.^26^ To evaluate residual confounding and selection biases, we assessed prespecified negative control outcomes (traumatic events [ICD-10-CM: S52.5, S52.6, S93.4, S43.0] and appendicitis [ICD-10-CM: K35]).^27^ To assess the robustness of the analytic approach, we repeated analyses in the full unmatched cohort, conducted additional stratified analyses by T2D subtype, and used Fine–Gray sub-distribution hazards models to account for competing risks.^28^ To assess whether differential screening intensity between individuals with and without T2D may have introduced surveillance bias, we restricted the breast, colorectal, and prostate cancer analyses to participants within recommended screening age ranges and adjusted for pre-index screening history (mammography, PSA testing, and colorectal cancer screening) in the prior five years (**Table S1**); Finally, to improve the specificity of the breast cancer outcome, we required confirmation of an ICD-10-CM–based diagnosis by the presence of at least one breast surgery CPT code within six months before or two years after the diagnosis date (**Table S1**).

All analyses were performed using R (version 4.5.1).

## RESULTS

The matched cohort consisted of 575,139 adults with newly diagnosed T2D and 689,719 matched adults without diabetes. Among adults with T2D, 40.7% were classified as MARD, 23.5% as MOD, 21.6% as SIDD, and 14.1% as the mixed subtype. After matching, age (64.7±12.3 [without diabetes] vs. 64.8±12.3 years [with T2D]), sex (50.3% female), and BMI (33.4±7.0 [without diabetes] vs. 33.3±6.7 kg/m² [with T2D]) were well balanced between groups, with standardized mean differences <0.1 (**Table 1**). Balance in comorbidity burden also improved after matching, with mean (SD) age-adjusted CCI values of 3.3 (1.6) in adults without diabetes and 3.5 (1.8) in adults with T2D. Pre-index healthcare utilization was similar between groups. Detailed descriptions of baseline characteristics of T2D subtypes are presented in **Table S3.**

**Table 1.**
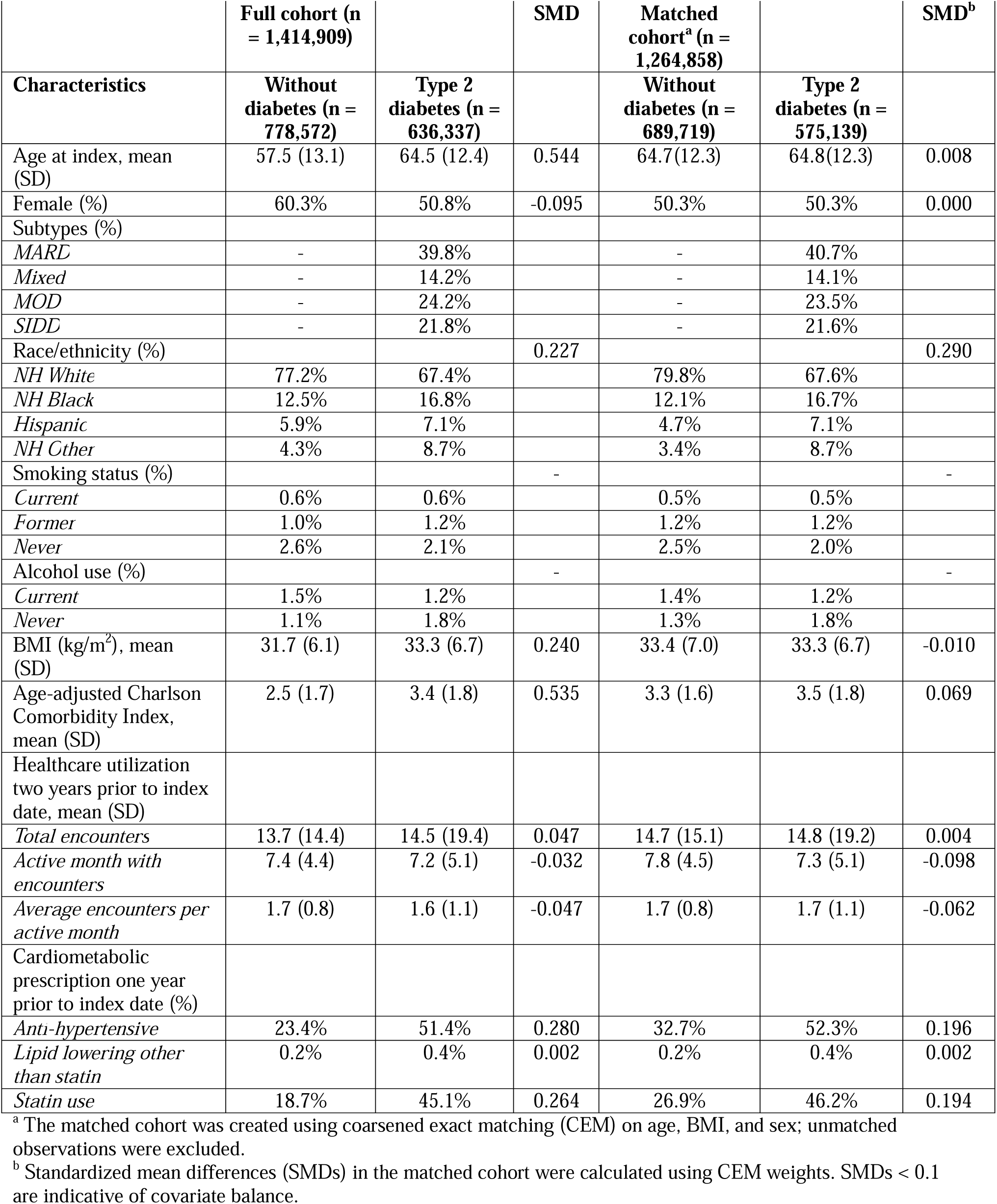
Baseline characteristics of adults with newly diagnosed type 2 diabetes and adults without diabetes in the full and matched cohorts.

|  | Full cohort (n = 1,414,909) |  | SMD | Matched cohort<sup>a</sup> (n = 1,264,858) |  | SMD<sup>b</sup> |
| --- | --- | --- | --- | --- | --- | --- |
| Characteristics | Without diabetes (n = 778,572) | Type 2 diabetes (n = 636,337) |  | Without diabetes (n = 689,719) | Type 2 diabetes (n = 575,139) |  |
| Age at index, mean (SD) | 57.5 (13.1) | 64.5 (12.4) | 0.544 | 64.7(12.3) | 64.8(12.3) | 0.008 |
| Female (%) | 60.3% | 50.8% | -0.095 | 50.3% | 50.3% | 0.000 |
| Subtypes (%) |  |  |  |  |  |  |
| *MARD* | - | 39.8% |  | - | 40.7% |  |
| *Mixed* | - | 14.2% |  | - | 14.1% |  |
| *MOD* | - | 24.2% |  | - | 23.5% |  |
| *SIDD* | - | 21.8% |  | - | 21.6% |  |
| Race/ethnicity (%) |  |  | 0.227 |  |  | 0.290 |
| *NH White* | 77.2% | 67.4% |  | 79.8% | 67.6% |  |
| *NH Black* | 12.5% | 16.8% |  | 12.1% | 16.7% |  |
| *Hispanic* | 5.9% | 7.1% |  | 4.7% | 7.1% |  |
| *NH Other* | 4.3% | 8.7% |  | 3.4% | 8.7% |  |
| Smoking status (%) |  |  | - |  |  | - |
| *Current* | 0.6% | 0.6% |  | 0.5% | 0.5% |  |
| *Former* | 1.0% | 1.2% |  | 1.2% | 1.2% |  |
| *Never* | 2.6% | 2.1% |  | 2.5% | 2.0% |  |
| Alcohol use (%) |  |  | - |  |  | - |
| *Current* | 1.5% | 1.2% |  | 1.4% | 1.2% |  |
| *Never* | 1.1% | 1.8% |  | 1.3% | 1.8% |  |
| BMI (kg/m<sup>2</sup>), mean (SD) | 31.7 (6.1) | 33.3 (6.7) | 0.240 | 33.4 (7.0) | 33.3 (6.7) | -0.010 |
| Age-adjusted Charlson Comorbidity Index, mean (SD) | 2.5 (1.7) | 3.4 (1.8) | 0.535 | 3.3 (1.6) | 3.5 (1.8) | 0.069 |
| Healthcare utilization two years prior to index date, mean (SD) |  |  |  |  |  |  |
| *Total encounters* | 13.7 (14.4) | 14.5 (19.4) | 0.047 | 14.7 (15.1) | 14.8 (19.2) | 0.004 |
| *Active month with encounters* | 7.4 (4.4) | 7.2 (5.1) | -0.032 | 7.8 (4.5) | 7.3 (5.1) | -0.098 |
| *Average encounters per active month* | 1.7 (0.8) | 1.6 (1.1) | -0.047 | 1.7 (0.8) | 1.7 (1.1) | -0.062 |
| Cardiometabolic prescription one year prior to index date (%) |  |  |  |  |  |  |
| *Anti-hypertensive* | 23.4% | 51.4% | 0.280 | 32.7% | 52.3% | 0.196 |
| *Lipid lowering other than statin* | 0.2% | 0.4% | 0.002 | 0.2% | 0.4% | 0.002 |
| *Statin use* | 18.7% | 45.1% | 0.264 | 26.9% | 46.2% | 0.194 |

<sup>a</sup> The matched cohort was created using coarsened exact matching (CEM) on age, BMI, and sex; unmatched observations were excluded.
<sup>b</sup> Standardized mean differences (SMDs) in the matched cohort were calculated using CEM weights. SMDs < 0.1 are indicative of covariate balance.

During a median follow-up period of 3.84 years (interquartile range [IQR], 1.75 – 6.59), 29,610 incident cancer cases were identified among adults with T2D (median follow-up time 3,41 [1.67-6.07]), and 31,730 cases were identified among adults without diabetes (median follow-up time: 4.33 [1.92-7.00]). Across all participants, prostate (8.68 per 1,000 persons) and breast (6.60 per 1,000 persons) cancers had the highest incidence rates, followed by colorectal cancer (1.56 per 1,000 persons).

Unadjusted cumulative incidence functions for site-specific cancers differed between T2D subtypes and individuals without diabetes (**Figure 2, Figure S2**). Across most cancer sites, cumulative incidence increased most rapidly among individuals with MARD and was generally lowest in MOD. For pancreatic and liver cancers, steeper increases were observed in the SIDD and Mixed subtypes compared with MOD and adults without diabetes. Prostate and breast cancer cumulative incidences were lower among SIDD and MOD relative to adults without diabetes. In contrast, individuals categorized as MARD and Mixed showed a cumulative incidence of these cancers comparable to or exceeding that of adults without diabetes.

**Figure 2.**
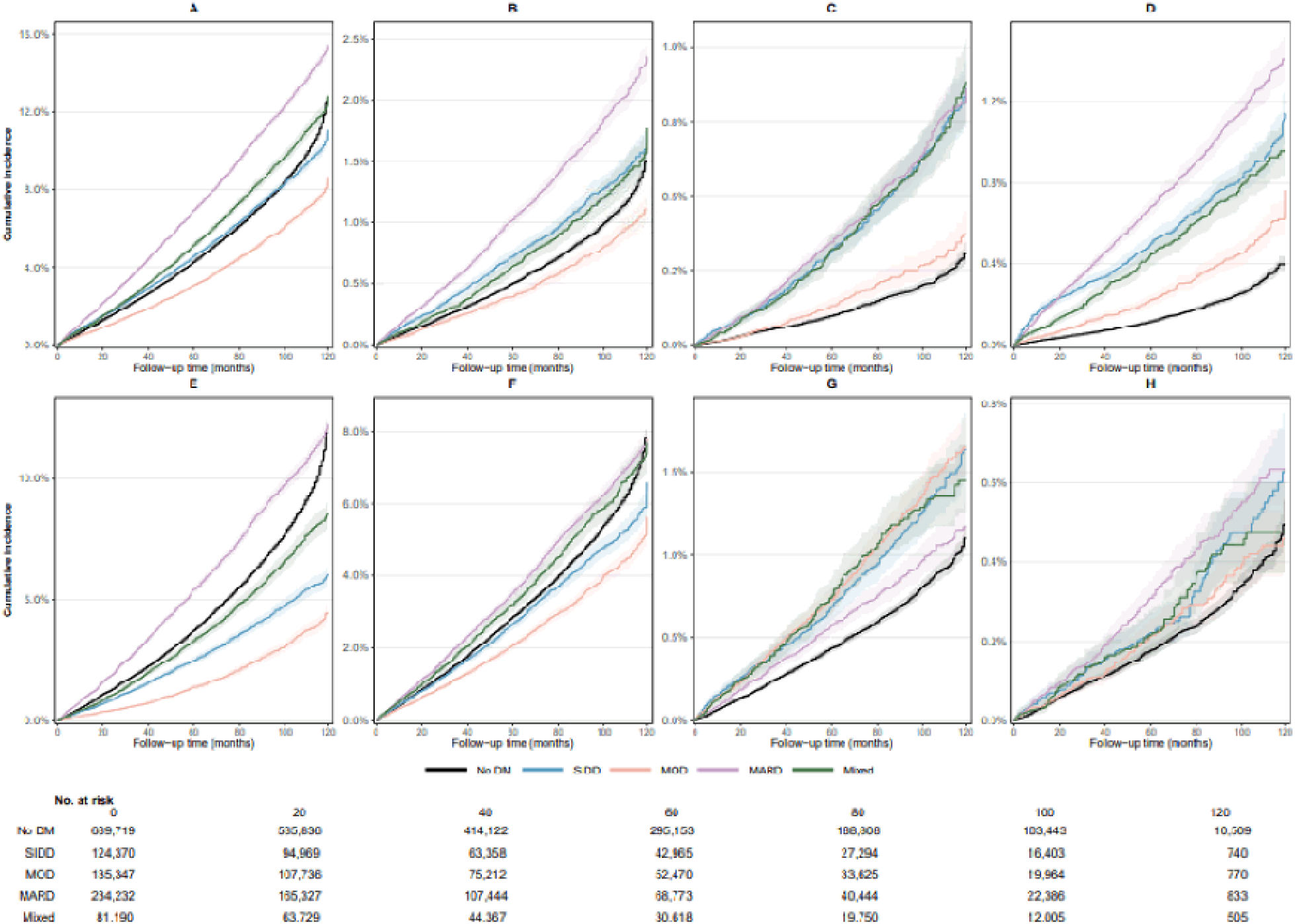
Cumulative incidence of cancer across type 2 diabetes subtypes and patients without diabetes. Cumulative incidence functions were estimated non-parametrically using the Aalen–Johansen estimator, accounting for competing events, and weighted using coarsened exact matching inverse probability weights. Curves are shown for individuals without diabetes (No DM) and for clinically defined subtypes (SIDD, MOD, MARD, and Mixed). Cancer sites include A) any cancer; B) colorectal cancer; C) liver cancer; D) pancreatic cancer; E) prostate cancer; F) breast cancer; G) endometrial cancer; and H) ovarian cancer. Time zero corresponds to the index date (diagnosis for cases and matched index date for no diabetes controls). Numbers at risk are shown at selected follow-up times. Curves for individuals without diabetes and with type 2 diabetes (not subtype-specific curves) are presented in **Figure S2**.

After covariate adjustment, pancreatic cancer hazards were elevated across all subtypes relative to adults without diabetes: SIDD (HR=3.87, 95% CI=3.51 to 4.27), Mixed (HR=3.16, 95% CI=2.81 to 3.57), MARD (HR=2.92; 95% CI=2.70 to 3.17), and MOD (HR=2.78, 95% CI=2.43 to 3.18) (**Figure 3**). Liver cancer hazards were similarly elevated across all subtypes, with the highest relative hazard in SIDD (HR=2.61, 95% CI=2.30 to 2.95). Colorectal cancer hazards were modestly elevated across all subtypes, with the strongest associations observed for SIDD (HR=1.43, 95% CI=1.33 to 1.54) and MARD (HR=1.40, 95% CI=1.33 to 1.48). Endometrial and ovarian cancer hazards were elevated across all subtypes (endometrial HRs, 1.19 to 1.41; ovarian HRs, 1.41 to 1.73).

**Figure 3.**
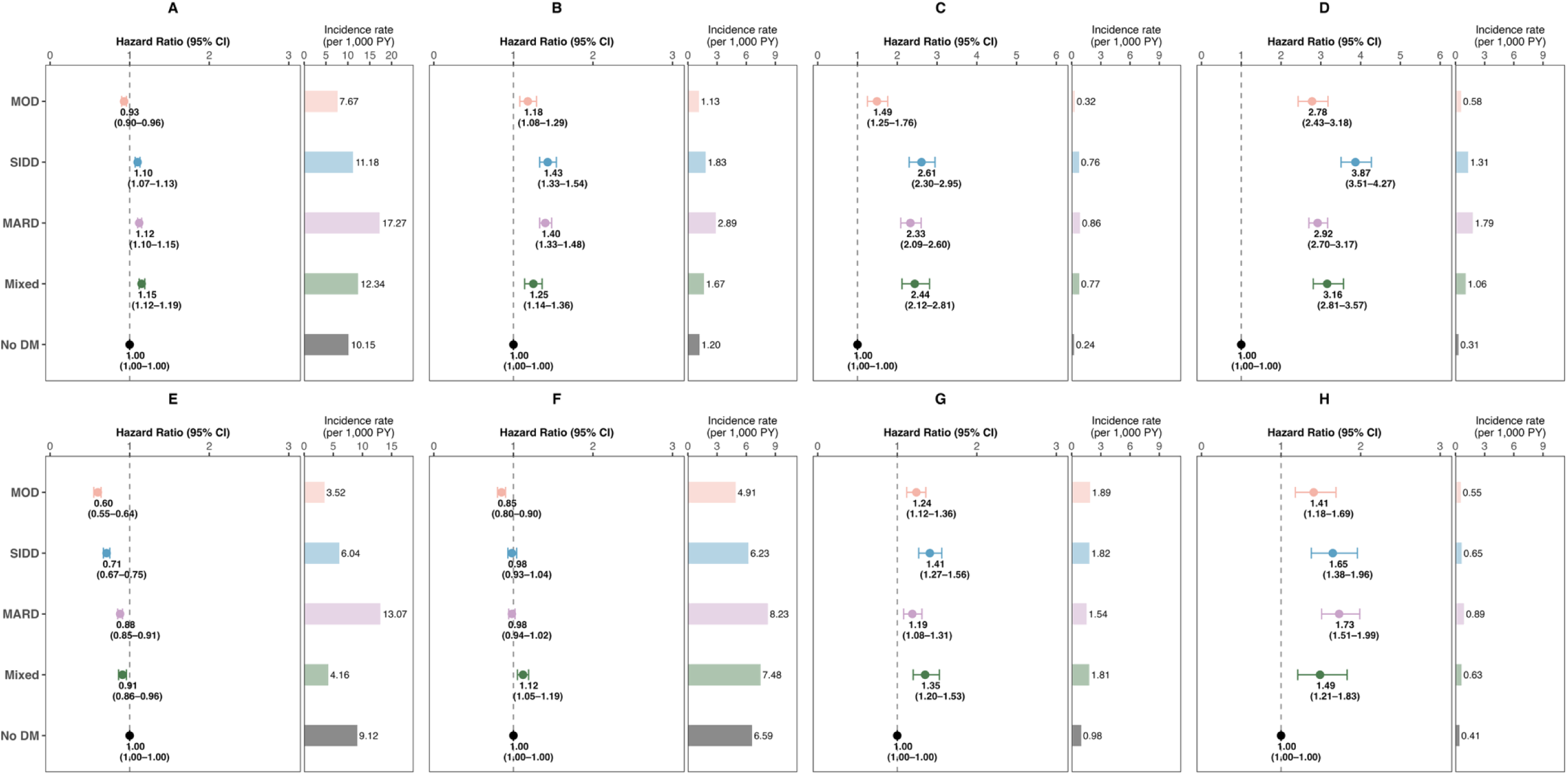
Site-specific cancer incidence across type 2 diabetes subtypes. Adjusted hazard ratios (left panels) and crude incidence rates per 1,000 person-years (right panels) are shown for individuals without diabetes and for clinically defined type 2 diabetes subtypes (SIDD, MOD, MARD, and Mixed). Cancer sites include A) any cancer; B) colorectal cancer; C) liver cancer; D) pancreatic cancer; E) prostate cancer; F) breast cancer; G) endometrial cancer; and H) ovarian cancer. Hazard ratios were estimated using cause-specific time-to-event regression models with no diabetes as the reference group, adjusting for demographic factors, healthcare utilization, and baseline clinical characteristics. Follow-up time was calculated from the index date (type 2 diabetes diagnosis for cases and matched index date for no diabetes comparators.

In contrast, prostate cancer hazards were lower across all subtypes relative to adults without diabetes, with the strongest inverse association in MOD (HR=0.60, 95% CI=0.55 to 0.64), followed by SIDD (HR=0.71, 95% CI=0.67 to 0.75), MARD (HR=0.88, 95% CI=0.85 to 0.91), and Mixed (HR=0.91, 95% CI=0.86 to 0.96). Breast cancer hazards showed a more heterogeneous pattern: MOD (HR=0.85, 95% CI=0.80 to 0.90), SIDD (HR=0.98, 95% CI=0.93 to 1.04), and MARD (HR=0.98, 95% CI=0.94–1.02) showed modestly lower hazards, while the estimate for Mixed was higher (HR=1.12, 95% CI=1.05–1.19) relative to adults without diabetes (**Figure 3**).

Screening uptake also differed across subtypes. Relative to individuals without diabetes, those with T2D had consistently lower uptake of mammography and PSA testing across all subtypes, with the largest differences observed in SIDD (mammography HR=0.75, 95% CI=0.74-0.76; PSA HR=0.72, 95% CI=0.70-0.75; **Table S4, Figure S3**). In contrast, colorectal cancer screening by stool-based testing was modestly higher among T2D subtypes, whereas visual inspection–based screening was broadly similar across groups.

Our findings were robust across the different sensitivity analyses. In lagged-time analysis, hazard ratio estimates were attenuated but remained directionally consistent for most cancer sites (**Table S5).** Negative control outcome analyses for traumatic injuries and appendicitis showed only small departures from the null (HRs of 1.04-1.27), indicating low residual confounding **(Table S6**). Analyses using multivariable regression without CEM matching, subtype stratification, and cause-specific Cox models yielded results comparable to those from the matched cohort (**Tables S7, S8, and S9**).

When analyses were restricted to screening-eligible age groups and adjusted for pre-index screening history, hazard ratio estimates for breast, colorectal, and prostate cancer were largely unchanged, suggesting that differential screening intensity before index date might not substantially confound the primary findings (**Table S4**). Lastly, after restricting breast cancer cases to ICD-10-CM diagnoses confirmed by at least one cancer treatment CPT code (57.6% of total cases), we observed stronger inverse associations across several T2D subtypes compared to those without diabetes (**Table S10**).

## DISCUSSION

In this large EHR-based matched cohort of over 1.2 million US adults, site-specific cancer rates and screening patterns varied across clinically defined subtypes of newly diagnosed T2D relative to adults without diabetes. Liver and pancreatic cancers consistently showed the strongest associations, particularly in individuals with SIDD and Mixed phenotypes, while prostate cancer rate was lower across all subtypes. Endometrial, ovarian, colorectal, and breast cancer rates exhibited more modest and variable patterns. This heterogeneity has direct implications for cancer risk stratification, surveillance, and prevention in diabetes care.

Large-scale observational studies and meta-analyses comprising over 32 million individuals reported 15-25% higher risks of cancer incidence and mortality in T2D.^14,29,30^ The highest excess risks have generally been reported for liver, pancreatic, and endometrial cancers, with more modest associations for breast, colorectal, and kidney cancers, whereas prostate cancer risk may be lower among men with T2D.^29–32^ Evidence on heterogeneity in cancer risk within T2D remains sparse. Prior work focused on diabetes timing or isolated metabolic traits: age at onset and prediabetes-to-diabetes transitions have been linked to modest differences in cancer risk, and glycemic burden and adverse lipid profiles have been associated with hepatopancreatic cancers through single-biomarker analyses.^15–18,33^ Our findings extend this literature by demonstrating that multidimensional phenotypes derived from routinely collected clinical features at diagnosis identify individuals with distinctly different site-specific cancer profiles.

The strongest associations were observed for liver and pancreatic cancers. In studies that treated T2D as a homogeneous exposure, reported relative risks ranged approximately 2.0–2.6 for liver cancer and 1.5–2.1 for pancreatic cancer.^14,29,30,32^ Our findings generally align with these estimates, though SIDD and Mixed subtypes were at the higher end of these ranges or above them. Early separation of cumulative incidence curves in these subtypes suggests that excess risk is established shortly after diabetes diagnosis rather than emerging only with longer disease duration, supporting the case for earlier risk stratification in metabolically high-risk subgroups.

These patterns are biologically plausible. SIDD is characterized by severe hyperglycemia, which may promote carcinogenesis through oxidative stress, DNA damage, and metabolic reprogramming.^34^ The Mixed subtype likely captures individuals with pronounced insulin resistance, for whom hyperinsulinemia and insulin–IGF signaling represent complementary carcinogenic mechanisms.^13^ Both pathways are especially relevant to liver and pancreas because these organs are central to glucose and lipid metabolism, suggesting potential convergency on shared tissue specific pathways that increase susceptibility.

Although relative hazards were often highest in SIDD, MARD contributed the greatest absolute cancer burden because it was the most prevalent subtype and was diagnosed at older ages, when baseline cancer risk is already high. The distinction between phenotype severity and phenotype prevalence has direct implications for how subtype information is applied in clinical versus public-health contexts.

Sex-specific cancers showed weaker and less coherent subtype patterning. Endometrial and ovarian cancers were elevated across several subtypes, with less separation than seen for hepatopancreatic cancer, suggesting shared hormonal and metabolic drivers rather than distinct subtype-specific mechanisms. Breast cancer showed the most heterogeneous patterns, with lower hazards in MOD, an elevated rate for Mixed, and near null estimates in other subtypes. This differs from several studies reporting positive associations between T2D and breast cancer when diabetes is treated as a homogeneous exposure.^14,32^ Although adiposity promotes postmenopausal breast cancer through increased estrogen synthesis, these findings suggest that the broader metabolic context, particularly the degree of insulin resistance, modifies susceptibility beyond what adiposity alone explains.

Prostate cancer showed the most consistent inverse associations, strongest in MOD. Prior cohort studies and meta-analyses have similarly reported lower prostate cancer risk in men with T2D, though estimates have varied across settings.^29,30,32^ A plausible explanation is detection bias related to PSA screening: higher BMI is associated with lower circulating PSA concentrations through hemodilution and hormonal alterations, reducing the likelihood of screening-detected disease.^35^ The lower PSA testing observed across T2D subtypes supports a role for reduced detection, and the strongest inverse association between prostate cancer and MOD, the subtype most characterized by obesity and adiposity, is consistent with this mechanism.^36^ Mammography uptake was likewise lower across all subtypes relative to adults without diabetes, while colorectal screening was broadly comparable, consistent with meta-analytic evidence.^38^ Collectively, detection bias likely contributes to weaker or inverse associations for breast and prostate cancer, while preserved colorectal screening rates strengthen the causal interpretation for elevated colorectal cancer risk across T2D subtypes.

Recent work has shown that routinely collected clinical variables can be used to optimize glucose-lowering treatments, and trial-based studies suggest treatment response differs across pathophysiological profiles.^39^ A parallel framework could be applied to cancer in T2D, in which clinically defined subtypes at diagnosis inform targeted surveillance and prevention. This approach could be further refined in EHR-linked biobanks that integrate longitudinal clinical records with genetic data,^40^ enabling more precise cancer risk profiling across T2D subtypes.

This study has several strengths. The large, nationally representative EHR cohort enabled precise estimation of site-specific cancer risks across clinically defined T2D subtypes. The matched cohort design, which aligned participants on age, sex, and BMI while restricting to those with documented healthcare utilization, effectively reduced confounding from differential surveillance and care-seeking behavior, a persistent concern in EHR-based cancer research.

Unlike etiological research cohorts, which have their own selection processes and may not reflect the breadth of clinical populations, our validated subtype definitions derived from routinely collected clinical variables are directly transferable to real-world clinical settings. Because research cohorts and EHR-based studies are subject to different forms of selection and ascertainment bias, which are most consequential for screening-sensitive cancers such as breast and prostate, the scale and random selection underlying our matched design help mitigate these concerns and strengthen causal inference. Moreover, rigorous analytic strategies, including restriction to incident T2D to reduce immortal time bias, lag-time analyses to mitigate reverse causation, and negative control outcome analyses to assess residual confounding, further support the internal validity of our findings.

Several limitations warrant consideration. Severe insulin-resistant diabetes (SIRD) was not separately identified due to limited model discrimination (AUC: 0.62) and the absence of fasting insulin measures in routine care. Individuals with pronounced insulin resistance may therefore have been distributed across other subtypes, potentially attenuating subtype-specific differences. Additionally, T2D cases were identified using the SUPREME-DM algorithm, which does not distinguish latent autoimmune diabetes in adults (LADA) from type 2 diabetes.

Misclassification of LADA cases into the SIDD subtype, given their shared features of insulin deficiency and severe hyperglycemia, cannot be fully excluded, though the cohort entry restriction of age ≥35 years should substantially limit this concern because of lower prevalence of LADA at older ages. Cancer outcomes were ascertained using diagnosis codes and may be subject to misclassification or incomplete capture of care received outside participating health systems. Although we conducted sensitivity analyses incorporating treatment-related CPT codes to improve outcome specificity for selected cancers, this approach may have introduced additional detection-related bias. Moreover, residual confounding from unmeasured factors, including diet, physical activity, body fat distribution, and medication use and adherence, cannot be excluded, particularly for obesity-related cancers. Finally, this analysis was restricted to newly diagnosed T2D and did not assess changes in subtype classification or cancer risk over longer disease duration.

## CONCLUSION

In this large EHR-based cohort, site-specific cancer burden and screening patterns among individuals with newly diagnosed T2D varied by clinically defined subtypes, with metabolically high-risk subtypes bearing disproportionately elevated rates for specific cancers and others showing neutral or inverse associations. Treating T2D as a homogeneous exposure obscures this heterogeneity and may partly explain inconsistent diabetes–cancer associations across studies and populations. Incorporating clinically observable diabetes subtypes into epidemiologic and clinical frameworks, together with attention to differential screening uptake, may improve cancer risk stratification and inform more targeted surveillance and prevention strategies in this population.

## Ethics approval and consent to participate

This study was exempt from ethical approval as it involved secondary analysis of pre-existing, HIPAA-limited, de-identified data from the Epic Cosmos Research Platform, which does not constitute human subjects research.

## Data availability

Epic Cosmos access is available through institutional representatives of participating institutions and the Epic Cosmos team after completing certification requirements. **Code availability**: The code for the analysis is available on https://github.com/chroniq-lab/diabetes_subtypes_cancers.

## Conflicts of Interest

None declared

## Funding

The work was funded by the Winship Invest$ Winter 2024 grant and the Emory Global Diabetes Research Center (EGDRC) Doctoral Support Fund. Zhongyu Li was supported through the Predoctoral Fellowship in Value Assessment and Health Outcomes Research from the Pharmaceutical Research and Manufacturers of America Foundation (PhRMA Foundation).

## Author contributions

ZL and JSV conceptualised the study. ZL and JSV developed the analytic plan with inputs from CH and MKA. ZL led the data extraction and analysis, and wrote the first draft. All authors contributed to the interpretation of the data. ZL is the guarantor of this work, and, as such, had full access to all the data in the study and take responsibility for the integrity of the data and the accuracy of the data analysis. All authors approved the final version of the manuscript.

## Acknowledgements

We thank A. Tobarran (Office of Information Technology, Emory Healthcare) and G. Najarro (Georgia Clinical and Translational Science Alliance, Emory School of Medicine) for facilitating access to the Epic Cosmos platform, as well as K. Mika and S. Luken from the Epic Cosmos team for their support. We would also like to thank B.C. Salazar, J. Guo, K. O. Sanaka, P. Vellanki, D. Hua, and T. Hung of Emory University for their inputs and comments at different stages of the analysis.

## Abbreviations

EHR: Electronic health record
MARD: Mild age-related diabetes
MOD: Mild obesity-related diabetes
SIDD: Severe insulin-deficient diabetes
SIRD: Severe insulin-resistant diabetes

## Notes

### Competing Interest Statement

The authors have declared no competing interest.

