## supplementary materials for "Site-Specific Cancer Incidence among Clinical Subtypes of Newly Diagnosed Type 2 Diabetes in the United States"

### Supplementary Methods

*Data source:* To prevent re-identification in Epic Cosmos Research Platform, geographic resolution is limited to the state level, and encounter dates are shifted at the patient level while preserving temporal intervals.

*Identification of adults with type 2 diabetes:* The Surveillance, Prevention, and Management of Diabetes Mellitus (SUPREME-DM) computable phenotype identifies diabetes from structured EHR elements, including diagnosis codes, laboratory results, and glucose-lowering medication data, and has been described in multisite EHR-based surveillance work. The date of type 2 diabetes diagnosis was defined as the date on which the second SUPREME-DM criterion was met.

*Body mass index extraction for adults without diabetes:* we extracted body mass index (BMI) measures recorded between 2012 and 2024 and excluded implausible values (<12 or >60 kg/m²).

*Coarsened exact matching procedure:* Matching strata were defined by the coarsened covariates (i.e., age at index date in 5-year categories, sex, and BMI categories at 18.5, 25.0, 30.0, 35.0, and 40.0 kg/m²). Strata with no observations on either side of the comparison were dropped from the analytic sample, and weights were assigned to retain all eligible matched observations within the remaining strata.^23^

*Type 2 diabetes subtype classification algorithm:* Adults with newly diagnosed type 2 diabetes were classified using a previously described EHR-based approach that operationalizes adult-onset type 2 diabetes subtypes from routinely available clinical variables (age at diagnosis, BMI, HbA1c, blood pressure, and lipid measures). Classification models for severe insulin-deficient diabetes (SIDD), mild age-related diabetes (MARD), and mild obesity-related diabetes (MOD) demonstrated strong discrimination (area under the curve [AUC] range: 0.81–0.99). Individuals were assigned sequentially to SIDD, MARD, and MOD. Those not meeting criteria for these subtypes were categorized as Mixed, reflecting individuals with overlapping metabolic features who do not meet the probability threshold for any dominant subtype profile.

Descriptive statistics: Continuous variables were summarized as mean and standard deviation (SD) if normally distributed or median (25^th^ percentile, 75^th^ percentile) otherwise. Categorical variables were described using frequencies and percentages.

### Supplementary Table 1. Diagnostic and procedural codes for cancer outcomes, screening, and treatment

| Site-specific Cancer | ICD-10-CM for primary cancer outcomes | CPT codes for screening (breast, colorectal, and prostate cancers) | CPT codes for breast cancer treatment ^a^ |
| --- | --- | --- | --- |
| Colorectal Cancer/visual and stool-based screening | C18, C19, C20, D01.0, D01.2, D01.1 | 45378, 45380, 45381, 45384, 45385, 45330, 82270, 82274, 81528, 74263 | - |
| Liver Cancer | C22.0 | - | - |
| Pancreatic Cancer | C25 | - | - |
| Breast Cancer/Mammography | C50, D48.6, D49.3 | 77067, 77063 | 19301-19307, 19296-19298, 96401-96425, 77385, 77386, 77401-77404, 77406-77409, 77411-77414, 77416, 77418, 77421, 77767-777772 |
| Endometrial Cancer | C54, C55, D07.0 | - | - |
| Ovarian Cancer | C56 | - | - |
| Prostate Cancer/prostate-specific antigen screening | C61 | 84152, 84153, 84154 | - |

ICD-10-CM: International Classification of Diseases, Tenth Revision, Clinical Modification

CPT codes: Current Procedural Terminology codes

^a^ when the procedure date preceded the ICD-10-CM date, it was used as the event anchor.

### Supplementary Table 2. Pre-specified cancer site-specific covariates used in fully adjusted models

| Site-specific Cancer | Site specific covariates identified by ICD-10 codes |
| --- | --- |
| Colorectal Cancer | History of kidney transplant, acromegaly, familial adenomatous polyposis, lynch syndrome, inflammatory bowel disease, cystic fibrosis, orchiectomy, personal history of colonic polyps and primary sclerosing cholangitis. |
| Liver Cancer | Hepatitis C, hepatitis B, alpha-1-antitrypsin deficiency, acute intermittent porphyria, cirrhosis, and hereditary hemochromatosis. |
| Pancreatic Cancer | Ataxia telangiectasia, cystic fibrosis, Li-Fraumeni syndrome, familial adenomatous polyposis, lynch syndrome, and other chronic pancreatitis |
| Female Cancers | Polycystic ovary syndrome, endometrial hyperplasia, menopausal disorders, hysterectomy (exclude if present), endometriosis, infertility, ovarian cysts, and oophorectomy (exclude if present). |
| Breast Cancer | No additional covariate. |
| Endometrial Cancer | Polycystic ovary syndrome, endometrial hyperplasia, menopausal disorders, and hysterectomy (exclude if present). |
| Ovarian Cancer | Endometriosis, infertility, ovarian cysts, and oophorectomy (exclude if present). |
| Prostate Cancer | Benign prostatic hyperplasia, prostatitis, and lower urinary tract symptoms |

### Supplementary Table 3. Descriptive characteristics of newly detected type 2 diabetes subtypes in Epic Cosmos in the matched cohort

|  | MOD  (n = 135,347) | SIDD  (n = 124,370) | MARD  (n = 234,232) | Mixed  (n = 81,190) |
| --- | --- | --- | --- | --- |
| Age at detection of SUPREME-DM | 53.0 (7.9) | 60.8 (12.2) | 74.3 (8.0) | 62.9 (5.9) |
| Female | 56.0% | 48.8% | 49.1% | 46.9% |
| **Race & Ethnicity** |  |  |  |  |
| *NH White* | 62.3% | 63.0% | 72.6% | 68.7% |
| *NH Black* | 20.7% | 19.8% | 13.3% | 15.3% |
| *Hispanic* | 8.7% | 9.2% | 5.1% | 7.1% |
| *NH Other* | 8.2% | 8.0% | 9.1% | 9.0% |
| **Key Biomarkers** |  |  |  |  |
| HbA_1c_ (mmol/mol) | 59.9 (5.4) | 88.2 (15.7) | 58.2 (6.7) | 63.0 (4.5) |
| HbA_1c_ (%) | 6.8 (0.6) | 10.1 (1.8) | 6.6 (0.8) | 7.2 (0.5) |
| Body mass index | 38.8 (6.0) | 34.0 (6.8) | 29.6 (5.0) | 33.8 (5.2) |
| Systolic BP | 130.9 (14.7) | 132.6 (16.3) | 131.2 (15.7) | 129.9 (15.0) |
| Diastolic BP | 78.9 (9.6) | 76.2 (10.4) | 71.6 (9.5) | 75.0 (9.3) |
| LDL cholesterol (mmol/l) | 97.3 (37.3) | 96.2 (41.8) | 81.4 (34.8) | 84.6 (35.6) |
| HDL cholesterol (mmol/l) | 43.9 (13.0) | 43.0 (15.1) | 47.9 (15.0) | 42.3 (13.8) |
| Triglycerides (mmol/l) | 173.7 (93.5) | 185.9 (106.2) | 141.4 (73.0) | 184.3 (102.4) |
| TGL:HDL ratio | 4.5 (3.1) | 5.1 (4.4) | 3.4 (2.3) | 5.1 (4.0) |
| Age-adjusted Charlson Comorbidity Index, mean (SD) | 2.1 (1.3) | 3.0 (1.7) | 4.5 (1.6) | 3.2 (1.3) |
| Healthcare utilization two years prior to index date, mean (SD) |  |  |  |  |
| *Total encounters* | 14.8 (18.3) | 12.7 (16.8) | 16.1 (21.1) | 14.2 (17.8) |
| *Active month with encounters* | 7.4 (5.1) | 6.6 (4.9) | 7.7 (5.3) | 7.2 (5.0) |
| *Average encounters per active month* | 1.7 (1.0) | 1.6 (1.0) | 1.7 (1.1) | 1.6 (1.0) |
| Cardiometabolic prescription one year prior to index date (%) |  |  |  |  |
| *Anti-hypertensive* | 48.6% | 46.8% | 57.4% | 52.4% |
| *Lipid lowering other than statin* | 0.3% | 0.4% | 0.5% | 0.5% |
| *Statin use* | 41.0% | 37.6% | 52.7% | 49.7% |

### Supplementary Table 4. Association of type 2 diabetes subtypes with cancer incidence and screening uptake

|  | **Events^a^** | **Censored** | **No Diabetes** | **MOD** | **SIDD** | **MARD** | **Mixed** |
| --- | --- | --- | --- | --- | --- | --- | --- |
| Colorectal | 6,332 | 985,357 | Ref | 1.12 (1.02, 1.23) | 1.37 (1.26, 1.50) | 1.34 (1.25, 1.45) | 1.19 (1.09, 1.31) |
| Breast | 17,962 | 574,274 | Ref | 0.93 (0.88, 0.99) | 0.99 (0.93, 1.05) | 0.91 (0.87, 0.97) | 1.04 (0.97, 1.10) |
| Prostate | 10,229 | 233,049 | Ref | 0.79 (0.72, 0.86) | 0.69 (0.64, 0.74) | 0.72 (0.68, 0.77) | 0.77 (0.72, 0.82) |
| Mammography | 333,320 | 258,306 | Ref | 0.84 (0.83, 0.85) | 0.75 (0.74, 0.76) | 0.82 (0.80, 0.83) | 0.85 (0.84. 0.87) |
| Protate specific antigen test | 66,425 | 176,853 | Ref | 0.83 (0.81, 0.86) | 0.72 (0.70, 0.75) | 0.81 (0.79, 0.83) | 0.75 (0.74, 0.77) |
| CRC visual test | 211,522 | 780,167 | Ref | 1.01 (1.00, 1.03) | 0.94 (0.93, 0.96) | 1.02 (1.00, 1,03) | 1.05 (1.03, 1.07) |
| CRC stool test | 56,836 | 934,853 | Ref | 1.11 (1.08, 1.15) | 1.25 (1.21, 1.28) | 1.20 (1.17, 1.24) | 1.15 (1.12, 1.19) |
| CRC any test | 249,981 | 741,708 | Ref | 1.03 (1.01, 1.04) | 0.99 (0.97, 1.00) | 1.05 (1.04, 1.07) | 1.06 (1.04, 1.08) |

^a^ estimates are from cause-specific Cox proportional hazards models adjusted for demographic factors, lifestyle factors, comorbidity, healthcare utilization, cardiometabolic medication use, cancer site-specific covariates, and pre-index screening history in the prior five years (see Methods).

### Supplementary Table 5. Association of type 2 diabetes subtypes with cancer outcomes after excluding diagnoses within two years (lagged analysis)

|  | **Events^a^** | **Censored** | **No Diabetes** | **MOD** | **SIDD** | **MARD** | **Mixed** |
| --- | --- | --- | --- | --- | --- | --- | --- |
| Colorectal | 4,916 | 1,252,432 | Ref | 0.99 (0.88, 1.11) | 1.31 (1.19, 1.45) | 1.17 (1.08, 1.26) | 1.12 (1.00, 1.26) |
| Liver | 1,524 | 1,258,541 | Ref | 1.48 (1.20, 1.81) | 2.91 (2.50, 3.39) | 2.51 (2.18, 2.87) | 2.61 (2.20, 3.10) |
| Pancreatic | 2,095 | 1,257,001 | Ref | 2.39 (2.02, 2.83) | 2.96 (2.58, 3.40) | 2.55 (2.28, 2.84) | 2.84 (2.43, 3.31) |
| Breast | 13,359 | 678,734 | Ref | 0.84 (0.78, 0.90) | 0.98 (0.91, 1.05) | 0.97 (0.92, 1.02) | 1.11 (1.03, 1.20) |
| Ovarian | 936 | 697,760 | Ref | 1.30 (1.02, 1.65) | 1.58 (1.25, 1.99) | 1.35 (1.11, 1.63) | 1.22 (0.92, 1.63) |
| Endometrial | 2,319 | 695,451 | Ref | 1.10 (0.97, 1.25) | 1.20 (1.04, 1.39) | 0.96 (0.84, 1.10) | 1.23 (1.04, 1.45) |
| Prostate | 13,425 | 540,842 | Ref | 0.60 (0.55, 0.65) | 0.69 (0.64, 0.74) | 0.85 (0.81, 0.88) | 0.91 (0.85, 0.97) |
| Any cancer | 38,431 | 1,199,552 | Ref | 0.87 (0.83, 0.91) | 1.02 (0.99, 1.06) | 1.04 (1.01, 1.07) | 1.11 (1.07, 1.16) |

^a^ estimates are from cause-specific Cox proportional hazards models adjusted for demographic factors, lifestyle factors, comorbidity, healthcare utilization, cardiometabolic medication use, and cancer site-specific covariates (see Methods).

### Supplementary Table 6. Association of type 2 diabetes subtypes with negative control outcomes

|  | **Events^a^** | **Censored** | **No Diabetes** | **MOD** | **SIDD** | **MARD** | **Mixed** |
| --- | --- | --- | --- | --- | --- | --- | --- |
| Appendicitis | 4,144 | 1,256,748 | Ref | 1.15 (1.03, 1.28) | 1.27 (1.14, 1.41) | 1.10 (1.00, 1.22) | 1.10 (0.97, 1.26) |
| Traumatic events | 16,160 | 1,244,732 | Ref | 1.06 (1.00, 1.12) | 1.06 (1.00, 1.13) | 1.07 (1.02, 1.12) | 1.04 (0.97, 1.11) |

^a^ estimates are from cause-specific Cox proportional hazards models adjusted for demographic factors, lifestyle factors, comorbidity, healthcare utilization, and cardiometabolic medication use (see Methods).

### Supplementary Table 7. Association of type 2 diabetes subtypes with cancer outcomes from the full cohort without matching

|  | **Events^a^** | **Censored** | **No Diabetes** | **MOD** | **SIDD** | **MARD** | **Mixed** |
| --- | --- | --- | --- | --- | --- | --- | --- |
| Colorectal | 9,274 | 1,405,635 | Ref | 1.24 (1.13, 1.35) | 1.49 (1.38, 1.60) | 1.45 (1.36, 1.53) | 1.30 (1.19, 1.42) |
| Liver | 2,517 | 1,412,392 | Ref | 1.63 (1.38, 1.94) | 2.77 (2.44, 3.15) | 2.34 (2.08, 2.63) | 2.58 (2.24, 2.98) |
| Pancreatic | 4,220 | 1,410,689 | Ref | 3.38 (2.94, 3.88) | 4.63 (4.18, 5.13) | 3.38 (3.08, 3.70) | 3.78 (3.33, 4.27) |
| Breast | 23,060 | 769,519 | Ref | 0.91 (0.86, 0.96) | 1.00 (0.95, 0.98) | 0.94 (0.90, 0.98) | 1.14 (1.07, 1.20) |
| Ovarian | 1,828 | 790,751 | Ref | 1.53 (1.28, 1.82) | 1.70 (1.43, 2.02) | 1.69 (1.47, 1.95) | 1.53 (1.24, 1.88) |
| Endometrial | 4,550 | 788,029 | Ref | 1.39 (1.26, 1.53) | 1.52 (1.37, 1.69) | 1.17 (1.06, 1.29) | 1.42 (1.26, 1.61) |
| Prostate | 22,345 | 599,027 | Ref | 0.67 (0.63, 0.72) | 0.72 (0.69, 0.76) | 0.82 (0.79, 0.85) | 0.94 (0.89, 0.99) |
| Any cancer | 67,515 | 1,347,394 | Ref | 0.99 (0.96, 1.03) | 1.13 (1.10, 1.16) | 1.10 (1.08, 1.13) | 1.19 (1.15, 1.23) |

^a^ estimates are from cause-specific Cox proportional hazards models adjusted for demographic factors, lifestyle factors, comorbidity, healthcare utilization, cardiometabolic medication use, and cancer site-specific covariates (see Methods).

### Supplementary Table 8. Association of type 2 diabetes subtypes with cancer outcomes from the primary models stratified by type 2 diabetes subtypes

|  | **Events^a, b^** | **Censored** | **No Diabetes** | **MOD** | **SIDD** | **MARD** | **Mixed** |
| --- | --- | --- | --- | --- | --- | --- | --- |
| Colorectal | 670/936/2,508/591 | 1,256,187 | Ref | 1.05 (0.94, 1.18) | 1.41 (1.28, 1.56) | 1.43 (1.35, 1.52) | 1.33 (1.18, 1.51) |
| Liver | 194/391/748/273 | 1,261,418 | Ref | 1.65 (1.31, 2.08) | 2.67 (2.21, 3.23) | 2.35 (2.06, 2.68) | 2.14 (1.74, 2.64) |
| Pancreatic | 342/670/1,554/376 | 1,260,097 | Ref | 2.95 (2.40, 3.62) | 3.95 (3.37, 4.63) | 2.95 (2.68, 3.24) | 2.95 (2.42, 3.60) |
| Breast | 1,647/1587/3,632/1,267 | 683,370 | Ref | 0.93 (0.87, 1.00) | 1.00 (0.93, 1.07) | 1.01 (0.96, 1.05) | 1.00 (0.93, 1.08) |
| Ovarian | 184/168/394/107 | 697,602 | Ref | 1.40 (1.12, 1.76) | 1.61 (1.27, 2.04) | 1.85 (1.58, 2.18) | 1.47 (1.10, 1.96) |
| Endometrial | 633/463/677/307 | 699,630 | Ref | 1.30 (1.16, 1.46) | 1.45 (1.26, 1.67) | 1.24 (1.11, 1.38) | 1.24 (1.05, 1.47) |
| Prostate | 908/1548/5,539/1,471 | 544,911 | Ref | 0.74 (0.67, 0.80) | 0.74 (0.69, 0.79) | 0.86 (0.83, 0.89) | 0.83 (0.76, 0.87) |
| Any cancer | 4,556/5,714/14,949/4,382 | 1,211,232 | Ref | 1.01 (0.96, 1.05) | 1.12 (1.08, 1.16) | 1.12 (1.10, 1.15) | 1.07 (1.03, 1.12) |

^a^ events are subtype only in the order of MOD/SIDD/MARD/Mixed

^b^ estimates are from cause-specific Cox proportional hazards models adjusted for demographic factors, lifestyle factors, comorbidity, healthcare utilization, cardiometabolic medication use, and cancer site-specific covariates (see Methods).

### Supplementary Table 9. Association of type 2 diabetes subtypes with cancer outcomes from the Fine-Gray subdistribution hazard models

|  | **Events^a^** | **Censored/competing event** | **No Diabetes** | **MOD** | **SIDD** | **MARD** | **Mixed** |
| --- | --- | --- | --- | --- | --- | --- | --- |
| Colorectal | 8,460 | 1,252,432 | Ref | 1.00 (0.90, 1.10) | 1.26 (1.17, 1.37) | 1.12 (1.05, 1.19) | 1.08 (0.98, 1.19) |
| Liver | 2,351 | 1,258,541 | Ref | 1.52 (1.27, 1.82) | 2.63 (2.30, 3.00) | 2.03 (1.79, 2.30) | 2.41 (2.07, 2.80) |
| Pancreatic | 3,891 | 1,257,001 | Ref | 2.92 (2.53, 3.37) | 3.93 (3.52, 4.38) | 2.76 (2.50, 3.04) | 3.20 (2.80, 3.64) |
| Breast | 20,665 | 678,734 | Ref | 0.87(0.82, 0.92) | 0.93 (0.88, 0.99) | 0.88 (0.84, 0.92) | 1.08 (1.02, 1.15) |
| Ovarian | 1,639 | 697,760 | Ref | 1.23 (1.01, 1.49) | 1.39 (1.16, 1.68) | 1.22 (1.04, 1.43) | 1.19 (0.95, 1.50) |
| Endometrial | 3,948 | 695,451 | Ref | 1.20 (1.08, 1.33) | 1.30 (1.17, 1.46) | 0.91 (0.82, 1.02) | 1.22 (1.06, 1.39) |
| Prostate | 20,651 | 540,842 | Ref | 0.63 (0.58, 0.68) | 0.65 (0.62, 0.69) | 0.73 (0.70, 0.76) | 0.87 (0.82, 0.92) |

^a^ estimates are from Fine and Gray subdistribution models adjusted for demographic factors, lifestyle factors, comorbidity, healthcare utilization, cardiometabolic medication use, and cancer site-specific covariates (see Methods).

### Supplementary Table 10. Associations of type 2 diabetes with breast cancer cases confirmed with both ICD10 and CPT codes

|  | **Events^a^** | **Censored/competing event** | **No Diabetes** | **MOD** | **SIDD** | **MARD** | **Mixed** |
| --- | --- | --- | --- | --- | --- | --- | --- |
| Breast cancer | 11,925 | 689,824 | Ref | 0.80 (0.74, 0.87) | 0.82 (0.77, 0.89) | 0.85 (0.80, 0.90) | 1.07 (0.99, 1.16) |

^a^ estimates are from cause-specific Cox proportional hazards models adjusted for demographic factors, lifestyle factors, comorbidity, healthcare utilization, cardiometabolic medication use, and cancer site-specific covariates (see Methods)

### Supplementary Figure 1. Index dates of patients with and without type 2 diabetes

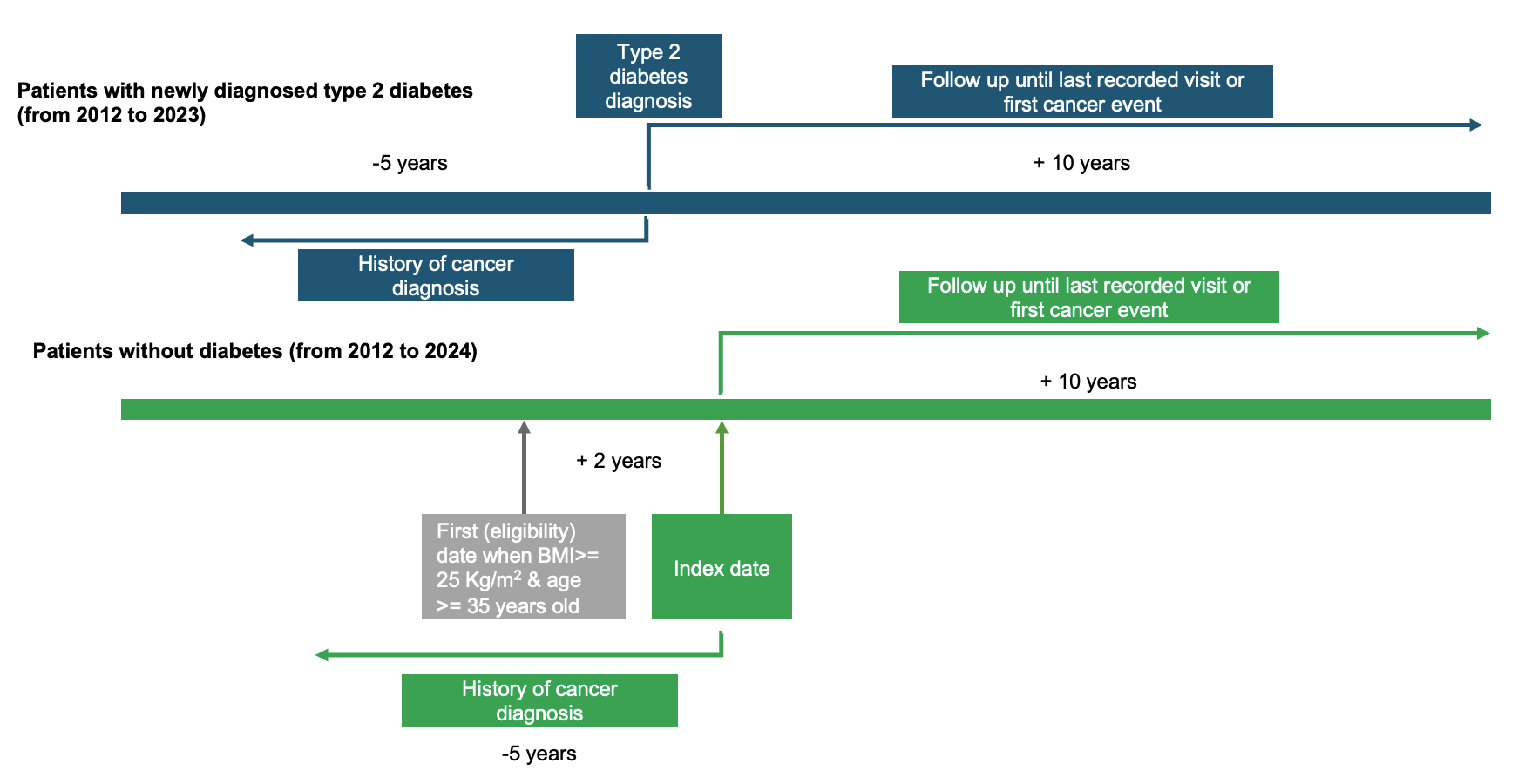

### Supplementary Figure 2. Cumulative incidence of cancer among patients with and without diabetes

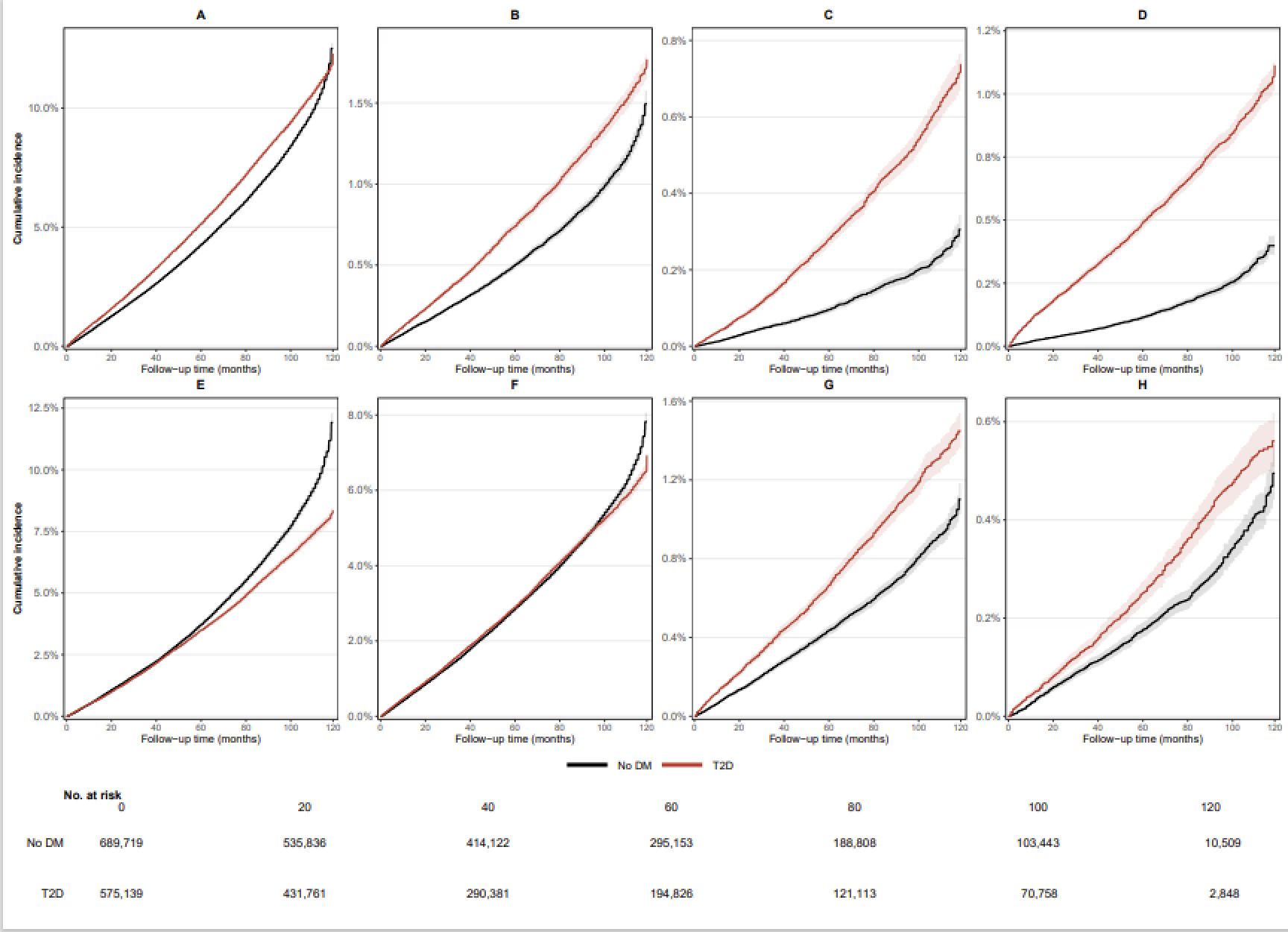

Cumulative incidence functions were estimated non-parametrically using the Aalen–Johansen estimator, accounting for competing events, and weighted using coarsened exact matching inverse probability weights. Curves are shown for individuals without diabetes and for individuals with type 2 diabetes (all subtypes combined). Cancer sites include: A) any cancer; B) colorectal cancer; C) liver cancer; D) pancreatic cancer; E) prostate cancer; F) breast cancer; G) endometrial cancer; and H) ovarian cancer. Time zero corresponds to the index date (type 2 diabetes diagnosis for cases and matched index date for no DM controls). Numbers at risk are shown at selected follow-up times.

### Supplementary Figure 3. Cumulative incidence of breast, prostate, and colorectal cancers and their screening tests

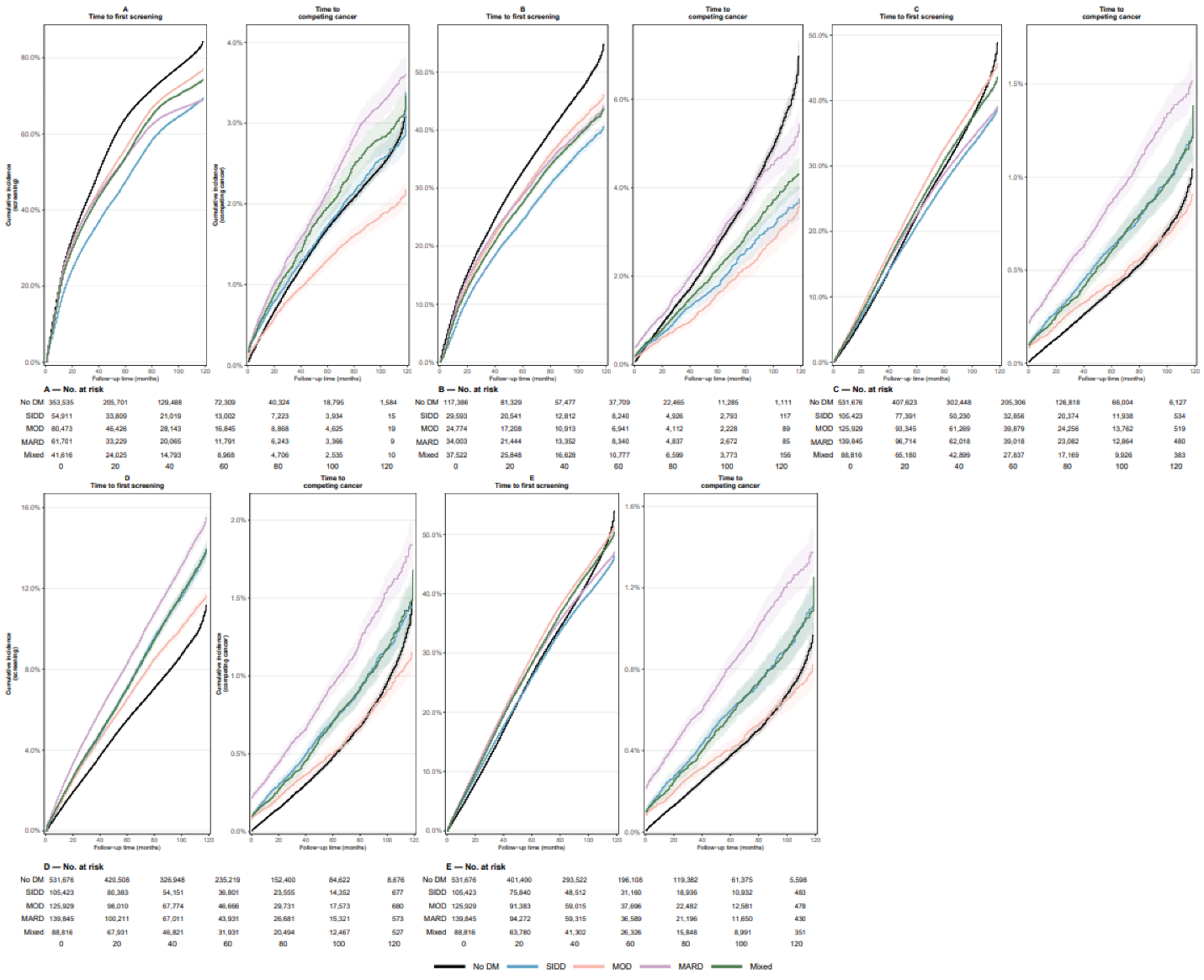

Cumulative incidence functions were estimated non-parametrically using the Aalen–Johansen estimator, accounting for competing events, and weighted using coarsened exact matching inverse probability weights. Curves are shown for individuals without diabetes and for clinically defined type 2 diabetes subtypes (SIDD, MOD, MARD, and Mixed). For each screening modality, the left panel shows time to first screening (event of interest) and the right panel shows time to first cancer diagnosis (competing event). Screening modalities include: A) mammography (women aged 40–74); B) PSA testing (men aged 55–69); C) colonoscopy/sigmoidoscopy (ages 45–75); D) stool-based CRC screening (ages 45–75); and E) any CRC screening (ages 45–75). Time zero corresponds to the index date. Numbers at risk are shown at selected follow-up times. Shaded bands indicate 95% confidence intervals.
